# Nucleocapsid antibody positivity as a marker of past SARS-CoV-2 infection in population serosurveillance studies: impact of variant, vaccination, and choice of assay cut-off

**DOI:** 10.1101/2021.10.25.21264964

**Authors:** Heather J Whitaker, Charlotte Gower, Ashley D Otter, Ruth Simmons, Freja Kirsebom, Louise Letley, Catherine Quinot, Georgina Ireland, Ezra Linley, Sonia Ribeiro, Shamez Ladhani, Jamie Lopez-Bernal, Gayatri Amirthalingam, Mary E Ramsay, Kevin E Brown

## Abstract

Serological surveillance studies sometimes use presence of anti-nucleocapsid antibody as a marker of natural SARS-CoV-2 infection. We explore seroconversion rates and antibody levels following Alpha and Delta variant infections, and vaccine breakthrough infections. We find lower seroconversion rates particularly following Alpha-variant vaccine breakthrough infections. We re-evaluate assay performance with a mix of past waned infections and recent breakthrough infections, that is relevant to current serological surveillance.

## Introduction

We explore the continued use of anti-nucleocapsid (N) antibody positivity as a marker of SARS-CoV-2 infections in serological surveillance studies following the introduction of vaccination and circulation of variants of concern. Recently, Allen et al.[1] described data for 23 study participants who experienced a breakthrough infection 14+ days after their 2^nd^ vaccine dose, of whom only 6 had detectable N antibodies 9-67 days post infection. If the sensitivity of the N assay is low for breakthrough infections, this has important implications for the interpretation of serological surveillance in a highly vaccinated population, such as UK blood donors [2]. We describe and compare the distribution and seropositivity of convalescent N antibody results in vaccinated and unvaccinated individuals, following Alpha or Delta variant infections.

### N antibodies following Alpha and Delta infections by vaccination status

Since February 2021 Public Health England have contacted a random sample of vaccine-eligible individuals identified from community testing data who had tested PCR-positive for COVID-19. The variant of each infection was determined by sequencing or S gene target failure [3] where available or assumed to be Alpha if the PCR date was before 03/05/2021 or Delta if the PCR date was after 24/05/2021. A convalescent blood sample was requested typically 4-6 weeks after infection. Sera were analysed using the Roche Elecsys anti-spike (S) and nucleocapsid (N) assays [4,5].

Convalescent N antibody results were available for 472 PCR positive cases recruited into the study. Individuals experienced infection between 08/02/2021-10/07/2021, with convalescent blood samples collected up to 11/08/2021. 24 were identified as having experienced past infection, either through self-report or N-positive sera taken during acute infection. At the time of infection, 143 were unvaccinated, 176 had received a single dose 21+ days prior and 80 were ≥7 days fully vaccinated. Exclusion of those with infection <21 days post dose 1 or re-infection left 379 individuals for study. The median age was 46 (min 19, max 87). 279 infections were allocated as Alpha variant, 88 Delta, 3 other lineages and 9 unknown. Samples were taken 22-100 days post onset or PCR positive with median 39 days, allowing at least 3 weeks for seroconversion.

Multivariable regression on log N levels to estimate the geometric mean ratio of antibody levels, and robust Poisson regression on N seropositivity to approximate prevalence ratios (PR), were carried out, including terms for vaccination status x variant interaction, age group, sex and a smoothing spline for days since onset (PCR date if unknown). Results are given in Table 1, along with seropositivity and the median and interquartile range of levels.

**Table 1.**

|  | N seropositivity |  | N antibody levels (AU/ml) |  |
| --- | --- | --- | --- | --- |
|  | n pos / N (%pos) | prevalence ratio<br>(95% CI)* | median (IQR) | geometric mean<br>ratio (95% CI)* |
| Alpha |  |  |  |  |
| unvaccinated | 103 / 110 (94%) | 1 (ref) | 46.1 (13.2 - 104) | 1 (ref) |
| single dose | 125 / 142 (88%) | 0.93 (0.86 - 1.01) | 9.2 (3.1 - 31.6) | 0.24 (0.15 - 0.38) |
| fully vaccinated | 21 / 27 (78%) | 0.84 (0.68 - 1.03) | 6.3 (1.1 - 19) | 0.19 (0.09 - 0.39) |
| Delta |  |  |  |  |
| unvaccinated | 23 / 24 (96%) | 1 (ref) | 39 (14.3 - 94.6) | 1 (ref) |
| single dose | 20 / 20 (100%) | 1.02 (0.93 - 1.11) | 29.9 (5.5 - 42.8) | 0.52 (0.18 - 1.52) |
| fully vaccinated | 42 / 44 (95%) | 0.98 (0.88 - 1.09) | 12.7 (5 - 34.6) | 0.36 (0.15 - 0.86) |

\*Multiple variable regression model included terms for vaccination status x variant, age group, sex, time since onset or original PCR test (spline)

A marginal reduction in prevalence of detectable N antibody following an Alpha vaccine breakthrough infection compared with infection in unvaccinated was found (single dose PR = 0.93 [95% CI 0.86 - 1.01, p=0.08]; fully vaccinated PR=0.84 [95% CI 0.68 - 1.03, p=0.09]), but no similar reduction was found for Delta breakthrough infections (single dose PR=1.02 [95% CI 0.93 - 1.11, p=0.69]; fully vaccinated PR = 0.98 [95% CI 0.88 - 1.09, p = 0.68]).

We found a statistically significant reduction in geometric mean N antibody level following both single dose and full vaccination for both Alpha and Delta infections. This is evident in Figure 1, which depicts the distribution of Roche N levels by vaccination status and variant. Following Alpha infections, the lower tail of the distribution sitting below the assay cut-off visually has greater density among individuals who were fully vaccinated at the time of infection. For Delta, while the distribution of levels is shifted downward in breakthrough infections as compared with vaccine-absent infections, seropositivity appears unaffected.

**Figure 1.**
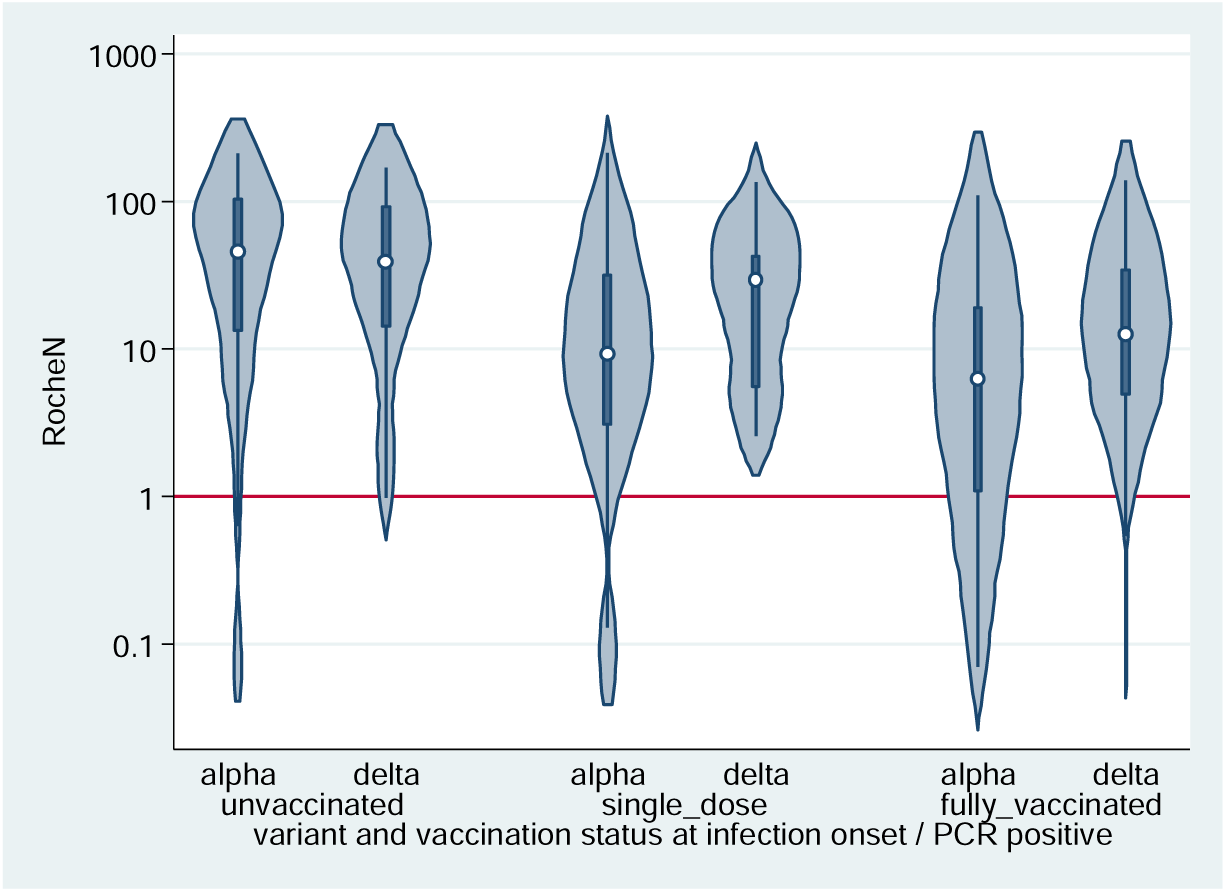
Violin plots depicting the smoothed distribution of Roche N levels (AU/ml) from convalescent sera. The white spot shows the median and the darker box the 25^th^ and 75^th^ percentiles. The horizontal red line is the assay positive/negative cut-off at 1.

### Optimal assay cut-offs for current serosurveillance

The Roche N assay cut-off of 1 was set high to give a clinical specificity of 99.8% [5]. We therefore explored an optimal cut-off of the Roche Elecsys anti-N assay by calculating the sensitivity, specificity and area under the receiver operating curve (ROC) for a range of values. To supplement the full 472 convalescent results described above, we included 220 Roche N results from secondary school pupils and staff with a record of testing PCR positive >21 days prior to sampling participating in the SKIDSplus study [6], and a panel of 233 convalescent individuals recruited during 2020, many of whom experienced infection early in the pandemic. We used only the most recent sample taken during 2021, thus our convalescent data includes a mix of samples from individuals who experienced wild-type SARS-CoV-2 in the past (max 431 days post infection), and more recent infections likely to be Alpha or Delta variant (primarily Alpha) across a broad range of ages. Two panels of pre-covid baseline sera were used: 472 residual from hospitals and labs and 995 Oxford biobank [7].

Of those ≥9 months post infection, 91% (106/116) were seropositive. A sensitivity of 97.2% was previously reported 20-82 days post wild-type infection [8], but waning will have led to reduced sensitivity [9].

Table 2 gives the sensitivity, specificity and area under ROC for a range of cut-offs. The manufacturers cut-off of 1 gives sensitivity 92.2% and specificity 99.9%. Area under ROC is maximised at 0.25, however the specificity of 99.0% at 0.25 seems lower than ideal. We consider the specificity of 99.6% at a cut-off of 0.4 to be preferable, which raises sensitivity to 94.1%.

**Table 2.** sensitivity, specificity, and area under the receiver operating curve (ROC) for a range of Roche Elecsys anti-nucleocapsid assay cut-offs

| Cut-off | sens | spec | ROC |
| --- | --- | --- | --- |
| 0.1 | 95.7% | 96.0% | 0.9585 |
| 0.15 | 95.5% | 98.3% | 0.9690 |
| 0.2 | 95.1% | 98.6% | 0.9686 |
| 0.25 | 94.9% | 99.0% | 0.9692 |
| 0.3 | 94.5% | 99.2% | 0.9686 |
| 0.35 | 94.3% | 99.4% | 0.9686 |
| 0.4 | 94.1% | 99.6% | 0.9685 |
| 0.45 | 94.0% | 99.6% | 0.9680 |
| 0.5 | 93.9% | 99.7% | 0.9681 |
| 0.55 | 93.8% | 99.7% | 0.9676 |
| 0.6 | 93.6% | 99.7% | 0.9665 |
| 0.65 | 92.8% | 99.7% | 0.9628 |
| 0.7 | 92.7% | 99.7% | 0.9622 |
| 0.75 | 92.7% | 99.7% | 0.9622 |
| 0.8 | 92.7% | 99.7% | 0.9622 |
| 0.85 | 92.7% | 99.8% | 0.9626 |
| 0.9 | 92.5% | 99.8% | 0.9615 |
| 0.95 | 92.3% | 99.9% | 0.9608 |
| 1 | 92.2% | 99.9% | 0.9602 |

Table 3 shows the percentage positive in our main data given cut-offs of 0.4 and 1. Notably, the percentage positive following Alpha infection rises by 1% in unvaccinated individuals and 2-3% in vaccinated individuals.

**Table 3:** number and percentage of Roche N levels (AU/ml) above thresholds of 0.4 and 1 in unvaccinated and vaccinated individuals following confirmed Alpha and Delta infections

| | Roche N level $\geq 0.4$<br>n / N (%) | Roche N level $\geq 1$<br>n / N (%) |
| --- | --- | --- |
| Alpha |  |  |
| unvaccinated | 105 / 110 (95%) | 103 / 110 (94%) |
| single dose | 128 / 142 (90%) | 125 / 142 (88%) |
| fully vaccinated | 22 / 27 (81%) | 21 / 27 (78%) |
| Delta |  |  |
| unvaccinated | 24 / 24 (100%) | 23 / 24 (96%) |
| single dose | 20 / 20 (100%) | 20 / 20 (100%) |
| fully vaccinated | 43 / 44 (98%) | 42 / 44 (95%) |

## Discussion

We conclude that N-antibody levels are lower following vaccination and differ by variant. Levels were generally higher following a Delta infection and seroconversion rates were unaffected by vaccination status at the time of infection. Even following Alpha infection for which levels were generally lower, 78% fully vaccinated individuals seroconverted. This is proportionally considerably higher than the 26% (6/23) found by Allen et al [1], however some sera were likely collected too soon to observe seroconversion (minimum 9 days), and 18 PCR positive individuals were asymptomatic, suggesting that seroconversion rates may be low following the very mildest infections. All Delta infections in our dataset were symptomatic, while 20/279 Alpha infections were asymptomatic. 65% (13/20) of these asymptomatic Alpha infections resulted in N seroconversion, compared with 91% (236/259) symptomatic Alpha infections (Fishers exact p=0.002).

Among those that did not seroconvert, ∼27% N levels were just below the threshold for positivity and we propose an equivocal range for the Roche Elecsys anti-N assay of 0.4-<1, which can be used to improve sensitivity while retaining a high specificity of 99.6%. Our convalescent data included mostly mild-moderate community past wild-type and more recent Alpha and Delta breakthrough infections, relevant to current serological surveillance. Sensitivity was estimated at 92.2% with the manufacturers cut-off of 1 or 94.1% using our proposed equivocal cut-off of 0.4.

Our findings mean that population estimates of N antibody seropositivity are likely to underestimate the proportion of the population who have experienced covid-19 in the highly-vaccinated UK population, particularly following an Alpha-variant wave or in younger populations who often experience mild infections, as well as due to waning. However, our data suggest that the majority do seroconvert, and that presence of N antibodies still serves as a reasonable marker for previous symptomatic infection regardless of vaccination status.

## Data Availability

Public Health England data requests were formerly made via the Public Health England Office for Data Release. At the time of submission this service is temporarily on pause while Public Health England transitions to the UK Health Security Agency.

## Ethnical statement

Serosurveillance of covid-19 is undertaken under Regulation 3 of The Health Service (Control of Patient Information) Regulations 2002 to collect confidential patient information (www.legislation.gov.uk/uksi/2002/1438/regulation/3/made) under Sections 3.(1)(b) 3.(1)(c), 3.(1)(d)(i-iii) and 3.(3)(a). The study protocols were subject to an internal review by the Public Health England Research Ethics and Governance Group (references NR0247, NR0228) and were found to be fully compliant with all regulatory requirements. As no regulatory issues were identified, and ethical review is not a requirement for this type of work, it was decided that a full ethical review would not be necessary.

## Acknowledgements

We acknowledge members of the PHE vaccine evaluation unit in handling and data management of the 2020 convalescent and SKIDSplus samples.

## Author contributions

HW carried out statistical analysis, with input on design and interpretation from KB, AO, HW, RS, GA, CG. HW wrote the manuscript, with input from CG, AO, GI, GA, KB. CG, FK, LL, CQ were responsible for the main sera collection and data management, led by JLB and MR. SL led the SKIDSplus study, GI, EL were responsible for data. GA and MR led the 2020 convalescent collection, data was managed by EL and SR. KB and AO oversaw sample testing.

## References

1. Allen N., Brady M., Carrion Martin A.I., Domegan L., Walsh C., Doherty L., Riain U.N., Bergin C., Fleming C., Conlon N. Serological markers of SARS-CoV-2 infection; anti-nucleocapsid antibody positivity may not be the ideal marker of natural infection in vaccinated individuals. Journal of Infection, 2021, ISSN 0163-4453, https://doi.org/10.1016/j.jinf.2021.08.012.

2. Whitaker H.J., Elgohari S., Rowe C., Otter A.D., Brooks T., Linley E. Impact of COVID-19 vaccination program on seroprevalence in blood donors in England, 2021. J Infect. 2021 S0163445321002243

3. Public Health England. Investigation of novel SARS-COV-2 variant: Variant of Concern 202012/01: Technical briefing document on novel SARS-CoV-2 variant. London: PHE; 2020. Available from: https://www.gov.uk/government/publications/investigation-of-novel-sars-cov-2-variant-variant-of-concern-20201201

4. Elecsys® Anti-SARS-CoV-2 S. Package Insert 2020-09, V1.0; Material Numbers 09289267190 and 09289275190. https://diagnostics.roche.com/gb/en/products/params/elecsys-anti-sars-cov-2-s.html

5. Elecsys® Anti-SARS-CoV-2. Package Insert 2020-07, V3.0; Material Numbers 09203095190 and 09203079190. F.Hoffman-La Roche https://diagnostics.roche.com/gb/en/products/params/elecsys-anti-sars-cov-2.html

6. Public Heath England. COVID-19 Surveillance in Children attending secondary schools. Protocol version 1.1, August 2020. Available at: https://assets.publishing.service.gov.uk/government/uploads/system/uploads/attachment_data/file/920906/sKIDsPLUS_protocol_v1_Aug20.pdf

7. The National SARS-CoV-2 Serology Assay Evaluation Group, The National SARS-CoV-2 Serology Assay Evaluation Group (2020): Head-to-head benchmark evaluation of the sensitivity and specificity of five immunoassays for SARS-CoV-2 serology on >1500 samples. figshare. Collection. https://doi.org/10.6084/m9.figshare.c.5046032.v1

8. Ainsworth M, Monique Andersson M, Auckland K, The National SARS-CoV-2 Serology Assay Evaluation Group. Performance characteristics of five immunoassays for SARS-CoV-2: a head-to-head benchmark comparison. The Lancet Infectious Diseases 2020, 20(12): 1390–1400. https://doi.org/10.1016/S1473-3099(20)30634-4

9. Harris RJ, Whitaker HJ, Andrews NJ, Aiano F, Amin-Chowdhury Z, Flood J, Borrow R, Linley E, Ahmad S, Stapley L, Hallis B, Amirthalingam G, Höschler K, Parker B, Horsley A, Brooks TJG, Brown KE, Ramsay ME, Ladhani SN. Serological surveillance of SARS-CoV-2: Six-month trends and antibody response in a cohort of public health workers. Journal of Infection 2021, 82(5): 162–169.

